# First-in-Human Safety and Tolerability Study of TOP-N53, a NO-Releasing PDE5 Inhibitor, in Healthy Volunteers

**DOI:** 10.64898/2026.04.15.26350931

**Authors:** Friedeborg Seitz, Hans Ulrich Gerth, Hermann Tenor, Christian Ludin, Yoshita Bhide, Matthias Schäfer, Jean-Luc Cracowski, Reto Naef

## Abstract

Chronic wounds, such as diabetic and ischemic ulcers, involve impaired perfusion and delayed healing. TOP-N53 is a novel bifunctional molecule combining nitric oxide (NO) release with phosphodiesterase-5 (PDE5) inhibition to enhance local NO–cGMP signalling, resulting in vasodilation and angiogenesis. This first-in-human, randomized, double-blind, vehicle-controlled Phase I trial assessed the safety, tolerability, pharmacokinetics (PK), and pharmacodynamics (PD) of single subcutaneous TOP-N53 doses in 29 healthy male volunteers. Each participant received injections of TOP-N53 and vehicle in the same forearm, but either at the proximal or at the distal site in an intra-individually blinded manner. Safety assessments included local and systemic parameters. PK and PD responses were evaluated by analysis of TOP-N53 and its bioactivation metabolite TOP-52 in plasma, and by Laser Speckle Contrast Imaging (LSCI), a non-invasive method to measure skin perfusion, respectively. TOP-N53 was safe and well tolerated, with no serious adverse events or local or systemic adverse reactions. Plasma concentrations remained below the quantification limit and LSCI showed sustained dose-dependent increases in local skin perfusion at doses of 4.84 µg and 9.075 µg TOP-N53 SC for up to 24 h post injection when compared to vehicle.

These findings support the favourable safety and tolerability profile of TOP-N53 associated with locally improved skin perfusion, encouraging its further clinical development as a topical treatment for chronic wounds with microvascular dysfunction.

## Introduction

Chronic wounds, including diabetic foot ulcers, venous leg ulcers, pressure ulcers, and ischemic skin lesions exemplified by digital ulcers in systemic sclerosis, remain a major clinical and socioeconomic burden worldwide. These wounds are characterized by delayed or incomplete healing due to persistent inflammation, hypoxia, impaired angiogenesis, and dysregulated matrix remodelling (1–3) . The underlying pathology is multifactorial, involving tissue ischemia, microbial colonization, oxidative stress, and a senescent wound-edge phenotype, which collectively inhibit re-epithelialization and closure (3–6). A key contributor impeding the closure of chronic cutaneous wounds and consequently, therapeutic target is compromised skin microcirculation largely governed by reduced microvascular density (capillary rarefaction) and an impaired ability to vasodilate (6).

Current therapeutic strategies for chronic wounds aim to control infection, promote tissue granulation, and restore local perfusion. Topical antimicrobials such as silver sulfadiazine or iodine-based agents are used to reduce bacterial load, while systemic antibiotics are reserved for clinically infected wounds (7). Enzymatic debriding agents like collagenase help remove necrotic tissue. Growth factors, including EGF, FGF, VEGF and GM-CSF have shown promise in clinical trials for diabetic foot ulcers and venous leg ulcers, but are not yet widely approved for clinical use. Topical growth factors such as recombinant human PDGF (becaplermin) stimulate proliferation and migration of wound fibroblasts, vascular smooth muscle cells, endothelial cells, and is FDA yet not EMA authorized for diabetic foot ulcers, though their efficacy is often limited by poor tissue penetration and high costs (8,9). Among systemic vasodilators, the endothelin antagonist bosentan, but neither macitentan nor ambrisentan, is recommended for the prevention of new digital ulcers in patients with systemic sclerosis, though hepatic dysfunction is a notable adverse effect requiring monitoring (10). However, bosentan is not effective for healing existing ulcers. Oral phosphodiesterase-5 (PDE5) inhibitors are recommended for digital ulcers (DU) healing in systemic sclerosis (SSc), exemplified by sildenafil (11) or tadalafil. Prostacyclin analogues such as intravenous iloprost (12), but not oral treprostinil are used as treatment for DUs in patients with SSc, at the cost of dose-dependent side effects (12). Consequently, oral PDE5 inhibitors and intravenous iloprost are recommended by the European Alliance of Associations for Rheumatology (EULAR) for this orphan condition (10). However, their use is associated with adverse reactions such as headache, dizziness, flushing, nausea, since high systemic exposure is required to obtain significant concentrations at site of ulcerations where capillary density is decreased (13). On the other hand, systemic vasodilators are currently not recommended for treatment of commonly occurring chronic wounds such as diabetic foot ulcer, pressure ulcer or venous leg ulcer. Adjunctive interventions like hyperbaric oxygen therapy can promote angiogenesis and healing but are often limited by logistical and economic barriers (14). Despite these options, many chronic wounds remain refractory to standard of care, underscoring the need for *on wound* administered, locally acting therapies aiming to reverse impaired wound perfusion without systemic adverse effects (15).

Nitric oxide (NO) plays a key role in orchestrating wound repair by regulating microcirculation vasodilation, angiogenesis, inflammation resolution, and antimicrobial defence (16–18). In chronic wounds, NO bioavailability is often diminished, resulting in impaired perfusion and delayed healing (18). Conversely, in early clinical trials *on wound* administered glycerol trinitrate releasing NO improved the low ulcer core perfusion of digital ulcers in systemic sclerosis (19) while a NO donating dressing device accelerated healing of diabetic foot ulcers (20). A hallmark among the multi-faceted roles of NO is its ability to stimulate the soluble guanylate cyclase, consequently, synthesis of cGMP supporting vasodilation and angiogenesis (21). In vascular cells, cGMP is mainly degraded by phosphodiesterase-5 (PDE5) and NO was described to activate PDE5, shortening its vasoactive efficacy (22). As a corollary, inhibition of PDE5 simultaneously to NO donors can be expected to prolong and amplify vasoactive effects emanating from the NO / cGMP pathway (22).

In a retrospective uncontrolled study, topical administration of the PDE5 inhibitor tadalafil (2% cream) on skin next to digital ulcers (DU) was correlated with benefit for healing of DU in patients with SSc, although the retrospective nature of the study prevented definitive conclusions from being drawn (23). In surgically created, full thickness, excisional cutaneous wounds of rabbits, *on wound* administered tadalafil in a hydrogel formulation dose-dependently accelerated wound healing (24). However, deficiency of NO from endothelial nitric oxide synthase (eNOS), the distinguishing feature of endothelial dysfunction occurring in diabetic foot ulcer and digital ulcers in systemic sclerosis limits efficacy of PDE5 inhibitors alone.

To cope with such environment, the NO-releasing PDE5 inhibitor TOP-N53 was developed as a locally applied, locally acting vasoactive treatment aiming to integrate high local efficacy to improve wound perfusion following its *on wound* administration with a minimal risk of systemic adverse reactions often seen with oral vasodilators (25). Following its intracellular (extrahepatic) bioactivation, the PDE5 inhibitor TOP-N53 is converted into nitric oxide (NO) and the even more potent PDE5 inhibitor TOP-52. The ensuing increase in cGMP synthesis (NO) and inhibition of its degradation by PDE5 (TOP-N53 and TOP-52) would synergistically enhance the cGMP pathway within the wound microenvironment and result in improved perfusion of chronic wounds (26, 27). Preclinical studies in murine models of diabetic, ischemic and normal wounds provided ample evidence that TOP-N53 significantly improves skin microvascular blood flow, promotes angiogenesis, and accelerates wound closure (26, 27).

To evaluate the clinical safety and tolerability of this therapeutic approach, a first-in-human, randomized, vehicle-controlled, double-blind (intraindividual), single ascending dose clinical trial was conducted in healthy male volunteers. The primary objective was to assess local safety and tolerability of a single subcutaneous injection of TOP-N53 or vehicle. Secondary objectives included evaluation of systemic safety parameters particularly systemic arterial blood pressure and pulse rate and characterization of the plasma pharmacokinetic profile of TOP-N53 and its (active) metabolite TOP-52. An exploratory objective was to assess changes in skin blood flow around the injection site using the non-contact Laser Speckle Contrast Imaging (LSCI) technique (28).

Findings from this study provide essential safety and tolerability data and support further clinical development of TOP-N53 as a localized treatment for digital ulcers in patients with systemic sclerosis or other chronic wounds.

## Methods

### Trial Design

This was a single-centre, randomized, double-blind (intra-individual), vehicle-controlled, first-in-human Phase I clinical trial conducted at CRS Clinical Research Services Mannheim GmbH (Germany) with TOPADUR Pharma AG acting as sponsor. The primary objective was to evaluate the local and systemic safety and tolerability of single ascending subcutaneous doses of TOP-N53 in healthy male volunteers. The study was conducted in full compliance with the principles of the Declaration of Helsinki and International Conference of Harmonization (ICH) Good Clinical Practice (GCP) guidelines as well as the Guideline on strategies to identify and mitigate risks for first-in-human and early clinical trials with investigational medicinal products - Revision 1 (EMEA/CHMP/SWP/28367/07 Rev. 1) and performed in accordance with EU CTR 536/2014.

The clinical study protocol (Version 2.0, dated 06 July 2020) received prior approval from both the Independent Ethics Committee (IEC) of the Landesärztekammer Baden-Württemberg (Chair: Prof. Dr. med. Gerlinde Egerer) and the German competent authority (Bundesinstitut für Arzneimittel und Medizinprodukte, BfArM). The IEC provided a favourable opinion on 16 July 2020, and regulatory approval was granted by the BfArM on 07 August 2020. All participants provided written informed consent prior to any study-related procedures. The trial was registered in the EudraCT database under number 2020-000672-38. The trial period was from 28-08-2020 (first participant first visit) to 02-07-2021 (last participant, last visit). A Clinical Study Report according to ICH E3 was compiled and archived (29).

### Participants

A total of 29 healthy male volunteers aged between 20 and 55 years were enrolled. Inclusion criteria included a body mass index (BMI) between 20.0 and 24.9 kg/m², Fitzpatrick skin phototypes I–IV and confirmed good general and in particular cardiovascular health based on comprehensive medical history, physical examination, and laboratory assessments. Key exclusion criteria were any history of dermatological conditions, systemic illness, recent use of medications, smoking, or participation in another clinical trial within the previous three months.

### Randomization, Blinding and Investigational Medicinal Products (IMP)

Participants were randomly assigned to receive a single subcutaneous dose of TOP-N53 into either the proximal (Area I) or distal (Area II) extensor side of the selected forearm and a matched placebo (vehicle) into either the distal (Area II) or proximal (Area I) extensor side of the same forearm (Fig. 1), in a double blinded manner, enabling intra-individual comparisons.

**Figure 1:**
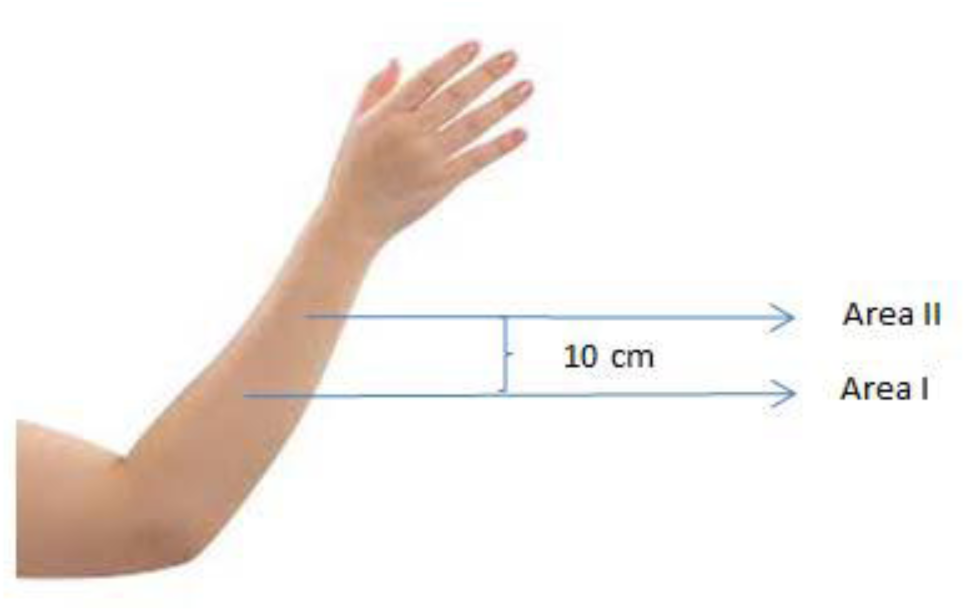
Injection areas. Area I: extensor side of the forearm near the elbow, Area II: extensor side of the forearm near the wrist. The distance between the proximal and distal injection site is 10 cm.

In this way, all participants received two SC injections, vehicle and TOP-N53. Neither the investigator nor the participant was aware whether the vehicle or TOP-N53 was injected at the distal or proximal injection site, but the investigator was aware of the dose level. The distance between the two injection sites was approximately 10 cm. A computer-generated simple randomization list was prepared and maintained by an independent pharmacist to ensure allocation concealment. Both, participants and investigators were blinded to treatment assignments.

Investigational Medicinal Products (IMP) were manufactured under GMP abiding by ICH and EMA quality guidelines. IMP Vehicle was composed of the following excipients: Cremophor EL at 1%, polyethylene glycol 400 at 10 %, phosphate buffered saline 89 %, all weight by weight, at pH 7.2. IMP TOP-N53 was a clear solution of TOP-N53 (10 µM, 6.05 µg / mL) in Vehicle. Dosing of TOP-N53 was by volume. Placebo consisted of IMP Vehicle alone. The trial included five ascending dose groups (DGs): 0.1 mL (dose group 1; vehicle or 0.605 µg TOP-N53), then 0.2 mL (dose group 2; vehicle or 1.21 µg TOP-N53), then 0.4 mL (vehicle or 2.42 µg TOP-N53), then 0.8 mL (vehicle or 4.84 µg TOP-N53), finally 1.5 mL (vehicle or 9.075 µg TOP-N53). One dose group (DG2) consisted of five participants; all others comprised six participants (Fig. 2).

**Figure 2:**
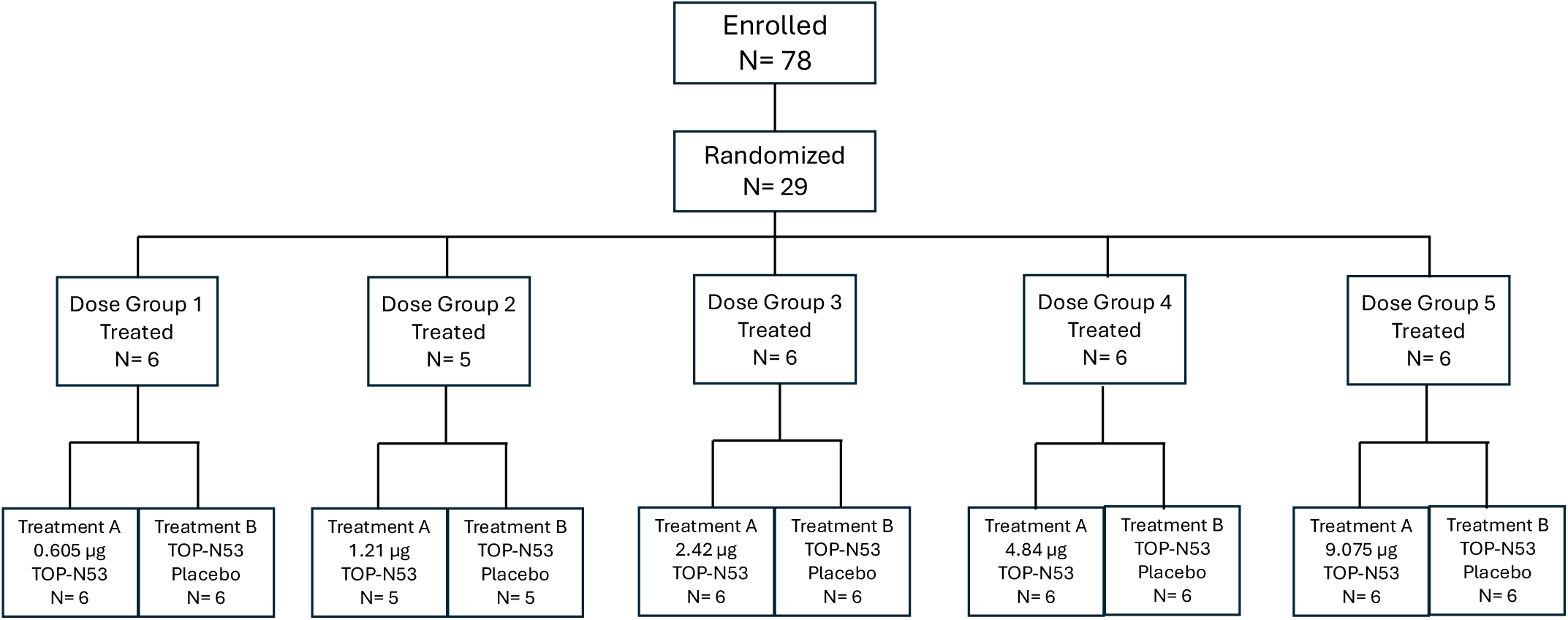
Disposition of participants and treatment assignments across the five dose groups.

In this trial, healthy volunteers who signed the informed consent form but before study specific procedures commenced were considered as enrolled participants. This explains that 78 participants were enrolled but 29 of those randomized with the remaining 49 excluded as screen failure because they did not meet all inclusion and exclusion criteria.

### Intervention

Dose escalation followed a sentinel dosing approach and was implemented in sequential cohorts. Progression to the next dose level occurred only after thorough review and clearance by an independent Data Safety Monitoring Board (DSMB). Injections were administered subcutaneously into predesignated areas of the forearm. To enable intra-individual control comparisons, each participant received both active treatment (TOP-N53 in vehicle) and placebo (vehicle) simultaneously, at identical volume.

### Safety and Tolerability Assessments

Safety assessments were the primary endpoints of the study and included both local and systemic evaluations. Local tolerability was assessed at each injection site through monitoring of erythema, swelling, pruritus, pain, and other dermal reactions. Systemic safety evaluations included regular monitoring of vital signs (blood pressure, heart rate, temperature), 12-lead electrocardiograms (ECGs), and comprehensive laboratory investigations (haematology, renal and hepatic panels). Adverse events (AEs) were recorded continuously during the 72-hour post-dose observation period.

### Pharmacokinetic and Pharmacodynamic Assessments

Blood samples were collected at baseline and at defined intervals up to 72 hours post-injection (i.e. at hours 0, 0.083, 0.25, 1, 2, 4, 6, 8, 24, 48, 72) to assess plasma concentrations of TOP-N53 and its primary metabolite, TOP-52. Quantification was performed using validated liquid chromatography–tandem mass spectrometry (LC-MS/MS) methods, with a lower limit of quantification (LLOQ) of 120 pg/mL for both TOP-N53 and TOP-52.

Pharmacodynamic (PD) effects were evaluated through Laser Speckle Contrast Imaging (LSCI), a non-contact, remote 2D imaging technique used to quantify cutaneous blood flow. LSCI measurements were recorded in regions of interests (ROI) surrounding the proximal (Area I) and distal (Area II) subcutaneous injection sites (Fig 1) at multiple time points (i.e. every 10 minutes during the first hour, every 20 minutes from the second to the fourth hour, every hour from the fifth to the eighth hour, then at 24 h) after IMP administrations. The measurements were performed in a calm environment with constant room temperature (air-conditioned) after an at least 15 min rest in supine position. The LSCI device was PeriCam PSI NR (Normal Resolution) from Perimed Järfälla, Sweden. The regions of interest (ROI) were defined circular areas around the injection sites, ROI 1 and ROI 2 of 15 mm diameter each for TOP-N53 or placebo (vehicle), and ROI 3 (about 5 mm in diameter) as reference area (located in the middle between ROI 1 and ROI 2 on the extensor side of the forearm). The device was positioned at approximately 30 cm distance from the skin surface; measurement duration was 1 minute. LSCI scans were done at the rate of one image per second in periods of 1 min.

The wavelength of the near infra-red (NIR) laser was 785 nm. The image size was 20×20 cm. Raw data computed by PIMSoft (Perimed) were arbitrary perfusion units (PU).

Skin blood flow was expressed as cutaneous vascular conductance (CVC). CVC was calculated as the flux expressed as PU divided by the mean arterial pressure (mm Hg) measured simultaneously once for every min period through an automated oscillometric device. By definition, the mean arterial pressure (MAP) was MAP = (SAP + 2xDAP)/3 where SAP and DAP are systemic and diastolic blood pressure, respectively. ΔCVC was calculated as the difference between CVC at a given time after IMP administration and immediately before, at baseline.

For the ΔCVC time course, Area Under the Effect Curves (AUEC), 0-1440 min after SC administrations, were computed for each individual treatment of a participant.

### Statistical Analysis

All statistical calculations (including ΔCVC AUEC) were carried out using SAS language and procedures (SAS version 9.4 SAS-Institute, Cary NC, USA).

The sample size was appropriate for a First-in-Human Phase I study aimed at preliminary safety and PD profiling. Safety, PK, and PD data were summarized using descriptive statistics.

No sample size calculation was done, and hypothesis testing was not planned. The trial was not powered to detect statistically significant differences between TOP-N53 and vehicle for the exploratory PD endpoint of skin perfusion.

*Post hoc*, the following statistical tests for comparisons of TOP-N53 versus vehicle on ΔCVC, MAP, and ΔCVC AUEC in Figure 4 were done using GraphPad Prism 10.4.1.

ΔCVC: 2-way ANOVA with Sidak’s multiple comparison test

MAP: 1-way ANOVA with Tukey’ s multiple comparison test (between any measurement time) ΔCVC AUEC: non-parametric Wilcoxon’s matched pairs signed rank test

## Results

### Participant Demographics

All 29 randomized male participants completed the study per protocol. Baseline characteristics, including age, body mass index (BMI), systemic systolic (SAP) and diastolic (DAP) arterial blood pressure, pulse rate and Fitzpatrick skin type, were well balanced across the five ascending dose groups. Participants ranged in age from 20 to 54 years, with a mean (± SD) age of 36.2 ± 11.15 years. BMI values fell within the inclusion criteria of 20.0–24.9 kg*m^-^², with a cohort mean of 23.21 ± 1.35 kg*m^-^². Systolic and diastolic systemic arterial pressure as well as pulse rate at the screening visit were also consistent with healthy population norms and the inclusion / exclusion criteria, and no significant differences were observed between groups (Table 1).

**Table 1:** Summary of demographic and anthropometric data at the screening visit for each dose group (DG1–DG5), with all participants being male. SAP, systolic systemic arterial blood pressure; DAP, diastolic systemic arterial blood pressure; SD, standard deviation.

| Dose Group (DG) | DG1 | DG2 | DG3 | DG4 | DG5 |
| --- | --- | --- | --- | --- | --- |
| TOP-N53 Dose (subcutaneous) | 0.605 $\mu\text{g}$ | 1.21 $\mu\text{g}$ | 2.42 $\mu\text{g}$ | 4.84 $\mu\text{g}$ | 9.075 $\mu\text{g}$ |
| Dose volume (mL) | 0.1 | 0.2 | 0.4 | 0.8 | 1.5 |
| Participants (n) | 6 | 5 | 6 | 6 | 6 |
| Age, Years; Mean (SD) | 39.0 (7.69) | 31.4 (14.69) | 38.2 (11.69) | 36.8 (12.19) | 34.8 (11.82) |
| BMI, $\text{kg}\cdot\text{m}^{-2}$; Mean (SD) | 24.08 (1.03) | 22.14 (1.46) | 22.83 (1.03) | 22.52 (1.28) | 23.28 (1.28) |
| SAP, mm Hg; Mean (SD) | 124.8 (10.4) | 126.8 (11.78) | 123.0 (6.23) | 125.7 (12.24) | 120.2 (7.49) |
| DAP, mm Hg; Mean (SD) | 77.3 (10.69) | 74 (10.91) | 78 (5.87) | 75 (12.74) | 68.8 (3.97) |
| Pulse Rate, $\text{min}^{-1}$; Mean (SD) | 64.5 (9.65) | 63.6 (8.44) | 56.5 (6.89) | 66 (10.97) | 59..5 (5.79) |

### Safety and Tolerability

There were no severe, serious, or fatal (SAE) adverse events, and none led to discontinuation. Overall, the number of treatment-emergent adverse events (TEAEs) was low, and none of the TEAE were considered as drug-related. Altogether, 5 TEAEs were observed in 5 of the 29 participants, with one event in Dose Group 2 (0.2 mL; 1.21 µg TOP-N53) and two each in Dose Group 3 (0.4 mL, 2.42 µg TOP-N53) and Dose Group 4 (0.8 mL, 4.84 µg TOP-N53). All TEAEs were of mild or moderate intensity and deemed unlikely or not related to study medication (IMP). Among the five TEAE there was 1 event of moderate dizziness (0.2 mL volume group) and 2 events of moderate headache (0.4 and 0.8 mL volume group).

The two TEAEs considered as unlikely related to the IMP were one event of moderate dizziness reported by one participant of Dose Group 2 (1.21 µg TOP-N53; 0.2 mL) and one event of moderate headache reported by one participant of Dose Group 3 (2.42 µg TOP-N53; 0.4 mL). Probably, dizziness was caused by the long fasting period after administration; the complaints were resolved immediately after food intake. During this episode, blood pressure and pulse rate values were within the normal ranges. The TEAE headache occurred at approximately 4.5 h after administration of the study drug and finally resolved after administration of two single doses of analgesic medication (ibuprofen). All other events were assessed as not related to the study drug (Table 2).

**Table 2:**
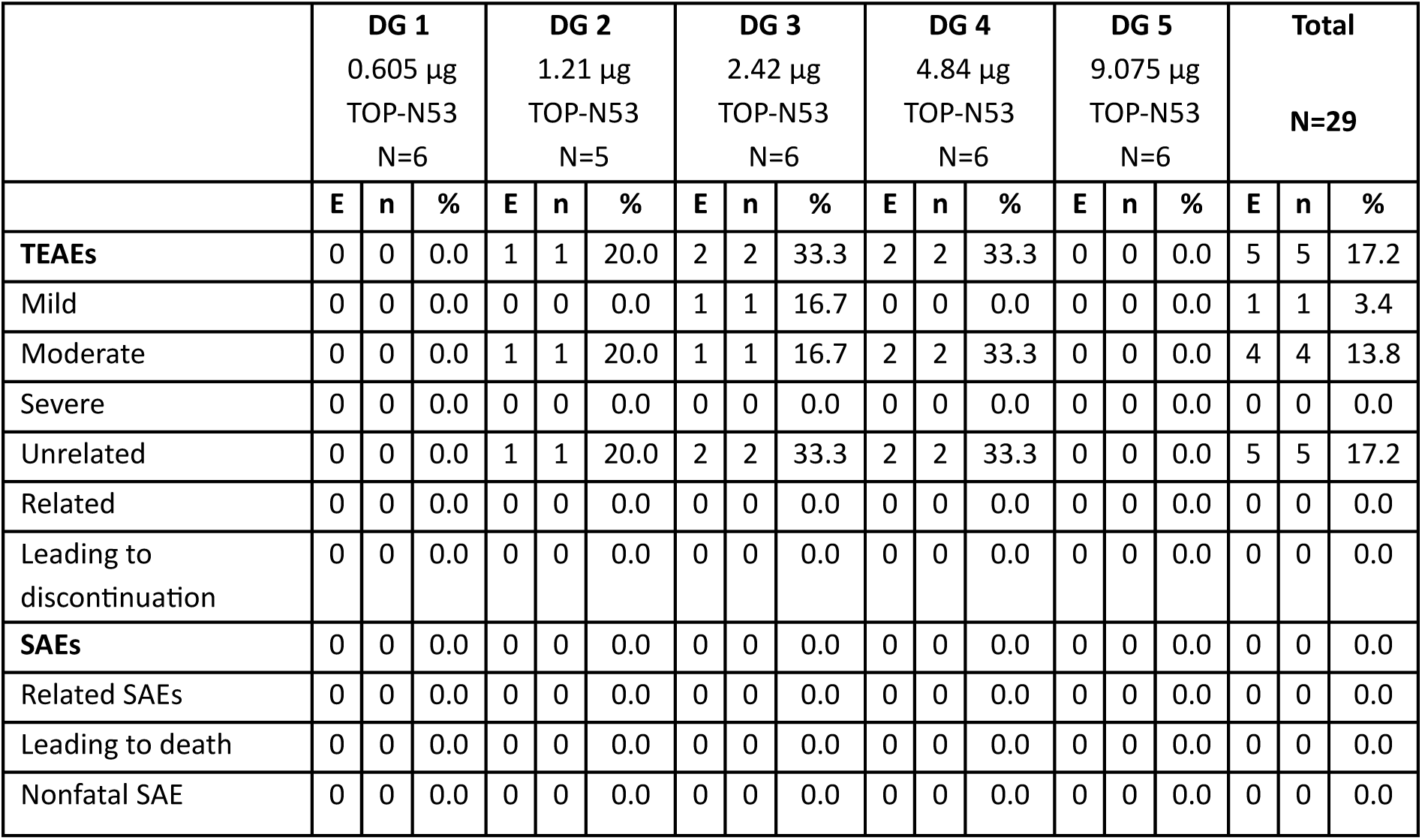
Overview of TEAEs and SAEs. DG: Dose Group, E: number of adverse events, N: number of participants in the respective group, n: number of participants with event, %: percent of participants with event, SAE: serious adverse event, TEAE: treatment-emergent adverse event. Adverse events that are not or unlikely related have been counted as unrelated, all other adverse events have been counted as related.

|  | DG 1<br>0.605 µg<br>TOP-N53<br>N=6 |  |  | DG 2<br>1.21 µg<br>TOP-N53<br>N=5 |  |  | DG 3<br>2.42 µg<br>TOP-N53<br>N=6 |  |  | DG 4<br>4.84 µg<br>TOP-N53<br>N=6 |  |  | DG 5<br>9.075 µg<br>TOP-N53<br>N=6 |  |  | Total<br>N=29 |  |  |
| --- | --- | --- | --- | --- | --- | --- | --- | --- | --- | --- | --- | --- | --- | --- | --- | --- | --- | --- |
|  | E | n | % | E | n | % | E | n | % | E | n | % | E | n | % | E | n | % |
| **TEAEs** | 0 | 0 | 0.0 | 1 | 1 | 20.0 | 2 | 2 | 33.3 | 2 | 2 | 33.3 | 0 | 0 | 0.0 | 5 | 5 | 17.2 |
| Mild | 0 | 0 | 0.0 | 0 | 0 | 0.0 | 1 | 1 | 16.7 | 0 | 0 | 0.0 | 0 | 0 | 0.0 | 1 | 1 | 3.4 |
| Moderate | 0 | 0 | 0.0 | 1 | 1 | 20.0 | 1 | 1 | 16.7 | 2 | 2 | 33.3 | 0 | 0 | 0.0 | 4 | 4 | 13.8 |
| Severe | 0 | 0 | 0.0 | 0 | 0 | 0.0 | 0 | 0 | 0.0 | 0 | 0 | 0.0 | 0 | 0 | 0.0 | 0 | 0 | 0.0 |
| Unrelated | 0 | 0 | 0.0 | 1 | 1 | 20.0 | 2 | 2 | 33.3 | 2 | 2 | 33.3 | 0 | 0 | 0.0 | 5 | 5 | 17.2 |
| Related | 0 | 0 | 0.0 | 0 | 0 | 0.0 | 0 | 0 | 0.0 | 0 | 0 | 0.0 | 0 | 0 | 0.0 | 0 | 0 | 0.0 |
| Leading to discontinuation | 0 | 0 | 0.0 | 0 | 0 | 0.0 | 0 | 0 | 0.0 | 0 | 0 | 0.0 | 0 | 0 | 0.0 | 0 | 0 | 0.0 |
| **SAEs** | 0 | 0 | 0.0 | 0 | 0 | 0.0 | 0 | 0 | 0.0 | 0 | 0 | 0.0 | 0 | 0 | 0.0 | 0 | 0 | 0.0 |
| Related SAEs | 0 | 0 | 0.0 | 0 | 0 | 0.0 | 0 | 0 | 0.0 | 0 | 0 | 0.0 | 0 | 0 | 0.0 | 0 | 0 | 0.0 |
| Leading to death | 0 | 0 | 0.0 | 0 | 0 | 0.0 | 0 | 0 | 0.0 | 0 | 0 | 0.0 | 0 | 0 | 0.0 | 0 | 0 | 0.0 |
| Nonfatal SAE | 0 | 0 | 0.0 | 0 | 0 | 0.0 | 0 | 0 | 0.0 | 0 | 0 | 0.0 | 0 | 0 | 0.0 | 0 | 0 | 0.0 |

No local reactions were observed (pain, erythema/redness, induration/oedema, haemorrhage/bruising or itching) at the injection site in any of the participants assigned to any of the IMP dose groups until 24 h after SC injection of TOP-N53 or placebo (vehicle).

Two participants reported delayed yet transient skin reactions (urticaria) at the vehicle (placebo) injection site, at 6 or 9 days after administration of study medication (IMP). The delayed skin reactions may have been associated with the vehicle used for SC administration of TOP-N53 and placebo.

No safety relevant changes on safety laboratory parameters (including standard haematology, coagulation, hepatic, renal, metabolism panels), vital signs (systemic arterial blood pressure, pulse rate), or ECG parameters were observed. Importantly, a loss in systemic arterial systolic or diastolic blood pressure was not observed up to and including 9.075 µg TOP-N53 (Fig. 3).

**Figure 3.**
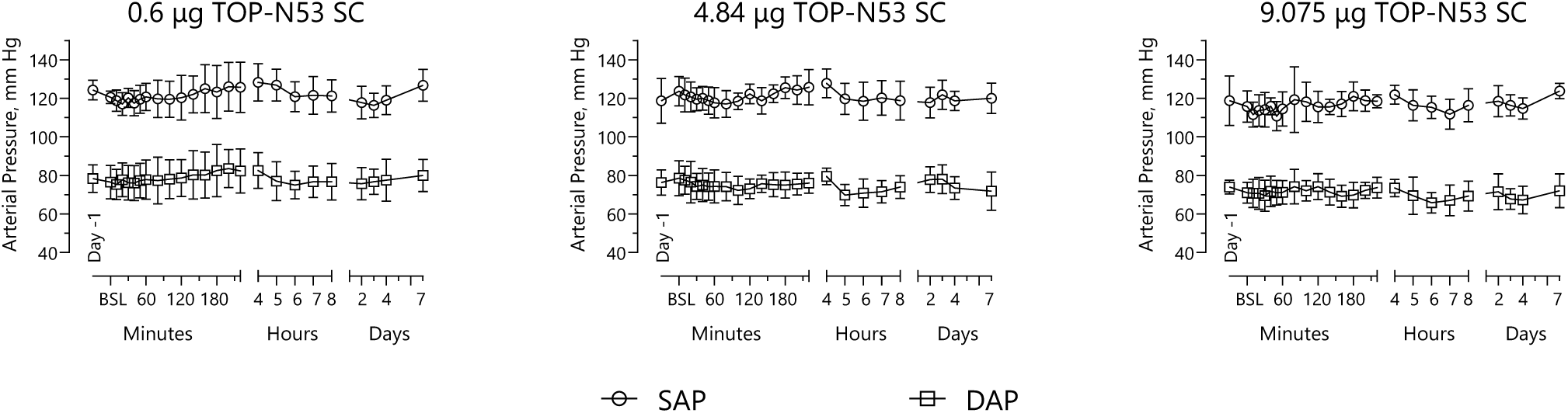
Effects of single-dose TOP-N53 (0.6 µg, 4.84 µg or 9.075 µg) on systolic (SAP) and diastolic (DAP) systemic arterial blood pressure. Measurements were done the day before IMP administration (day -1), at 10 min prior IMP administration (BSL) and then at the indicated times after IMP administration. Results are shown as the arithmetic means ± SD of n=6 participants for each dose group.

Overall tolerability at study end was rated as good for all participants. No clinically relevant abnormalities were attributed to the IMP (TOP-N53 or vehicle).

### Pharmacokinetics

Plasma concentrations of both TOP-N53 and its primary metabolite, TOP-52, were generally below their lower limit of quantification (LLOQ, 120 pg/mL), consistent with minimal systemic absorption. Of 29 participants, only two participants in the lowest dose group (0.1 mL) exhibited quantifiable plasma concentrations of TOP-N53 with 259.1 pg/mL at baseline and 400.6 pg/mL at 24 hours post-dose. These values were deemed analytically implausible and likely unrelated to study drug exposure. No quantifiable concentrations of TOP-52 were detected in any participant.

### Pharmacodynamics: LSCI measurements of local skin perfusion

Pharmacodynamic effects were evaluated by Laser Speckle Contrast Imaging (LSCI), quantifying skin blood flow around the SC injection sites expressed as changes of cutaneous vascular conductance (ΔCVC) over time after SC administration of TOP-N53 or vehicle (placebo) versus baseline (CVC prior treatment). A sustained increase in local skin perfusion was observed at 4.84 µg TOP-N53 versus vehicle, both SC in 800 µl volume (Fig 4J) and 9.075 µg TOP-N53 versus vehicle, both SC in 1500 µl volume (Fig 4M) and detectable at almost all measurements times after SC injection until the last assessment at 24 hours. This increase in local skin perfusion by TOP-N53 at 4.84 and 9.075 µg was deemed dose-dependent when compared to TOP-N53 at the lower doses of 0.6, 1.21 or 2.42 µg (Fig 4A, D, G).

**Figure 4.**
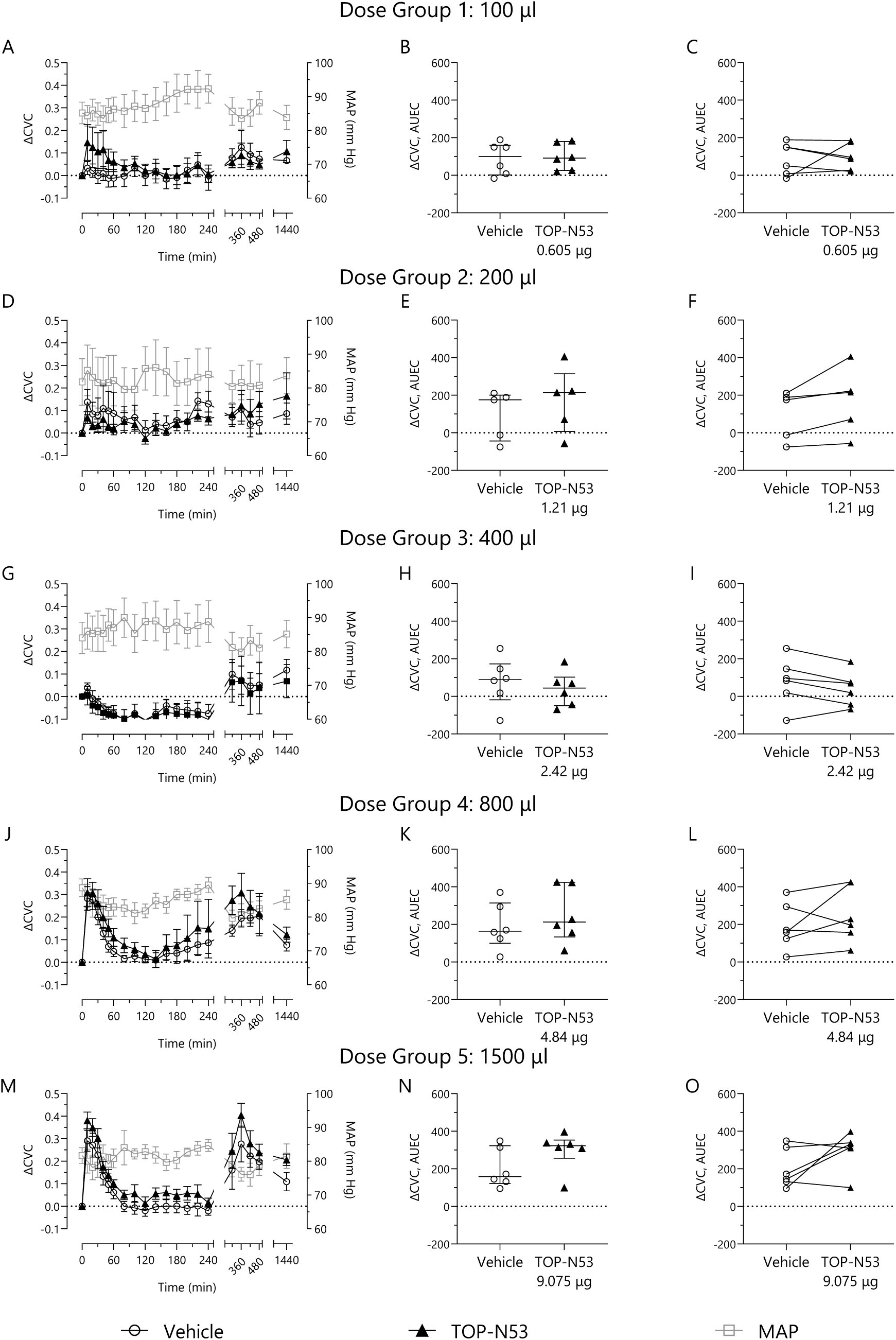
Effects of single-dose, subcutaneous administration of TOP-N53 at 0.605 µg (A,B, C), 1.21 µg (D, E, F), 2.42 µg (G, H, I), 4.84 µg (J, K, L) or 9.075 µg (M, N, O) on skin blood flow in healthy volunteers versus vehicle, measured by LSCI. Dosing was by volume from a solution of TOP-N53 in vehicle at 6.05 µg / ml (10 µM). The dose volume was identical for TOP-N53 and vehicle with 100 µl in A-C (Dose Group 1), 200 µl in D-F (Dose Group 2), 400 µl in G-I (Dose Group 3), 800 µl in J-L (Dose Group 4), 1500 µl in M-O (Dose Group 5). Perfusion units (PU) were normalized to mean arterial blood pressure (MAP) to calculate cutaneous vascular conductance (CVC). ΔCVC represents change from baseline. Left panels A, D, G, J, M show time course (0–24 h) of ΔCVC. Results from oscillometric MAP measurements run in parallel to LSCI are added. Data are shown as arithmetic means with SEM from n=6 participants for Dose Group 1, 3-5 and n=5 participants for Dose Group2. Middle panels B, E, H, K, N show computed values for ΔCVC Area Under the Effect Curves (AUEC, min * PU * mm Hg^-1^), 0-1440 min of individual participants. Median with the interquartile range (Q25, Q75) is shown. Right panels C, F, I, L, O illustrate pairwise comparisons of ΔCVC AUEC determined for vehicle and TOP-N53 in each participant. ΔCVC AUEC values of individual participants are indicated by symbols. p>0.05 for TOP-N53 versus vehicle at all time points (A, D, G, J, M) (2-way ANOVA with Sidak’s multiple comparison test) and for MAP (between any measurement times, 1-way ANOVA with Tukey’ s multiple comparison test). Differences between ΔCVC AUEC determined with vehicle or TOP-N53 were analysed using non-parametric Wilcoxon’s matched pairs signed rank test. No significant differences were found in any of the dose groups. All statistical tests were post hoc and computed by GraphPad Prism 10.4.1.

Values for Area Under the Effects Curves (AUEC) were calculated from the time course of ΔCVC for each individual participant in all dose groups separately for vehicle and TOP-N53. Results for dose group 1 to 5 are shown in Fig 4 B, E, H, K, N as individual data with the median and the interquartile range (Q25, Q75) and as pairwise comparisons, ΔCVC AUEC Vehicle versus ΔCVC AUEC TOP-N53 for each individual participant in Fig 4 C, F, I, L, O. These ΔCVC AUEC (0-1440 min) confirmed that for the two highest dose groups, in particular with TOP-N53 at 9.075 µg changes in cutaneous vascular conductance from baseline over the 24 hours period following its single, subcutaneous administration were more marked than with vehicle. On the other hand, in dose groups 1 and 3 with TOP-N53 at 0.605 µg and 2.42 µg there was no difference versus vehicle. In dose groups 4 and 5 pairwise comparisons showed that in 4 from 6 participants ΔCVC AUEC at the TOP-N53 (4.84 or 9.075 µg) SC injection site was higher than at the vehicle injection site (Fig 4 L and O), while in dose groups 1 and 3 the same occurred in only 2 or 1 participants, respectively (TOP-N53 at 0.605 or 2.42 µg, Fig 4C and I). In dose group 2 (200 µl volume SC) ΔCVC AUEC seemed higher with TOP-N53 (1.21 µg) versus vehicle (Fig 4E, Table 3), in pairwise comparisons for all participants (Fig 4F). However, ΔCVC was less with TOP-N53 (1.21 µg) than vehicle up to 6 hours after injection but conversely higher than vehicle at the remaining measurements at 7 h, 8 h and 24 h. Overall, the variability of ΔCVC AUEC was rather high (except for 5 of the 6 participants on TOP-N53, 9.075 µg in dose group 5).

**Table 3:** ΔCVC AUEC (min * PU * mm Hg^-1^) (0-1440 min after SC administration of vehicle or TOP-N53) of CVC changes versus baseline (ΔCVC) – Arithmetic means per dose group (DG) and treatment regimen (vehicle or TOP-N53 SC).

| Dose Groups (DG) | DG1: 100 $\mu\text{l}$<br>n=6 | | DG2: 200 $\mu\text{l}$<br>n=5 | | DG3: 400 $\mu\text{l}$<br>n=6 | | DG4: 800 $\mu\text{l}$<br>n=6 | | DG5: 1500 $\mu\text{l}$<br>n=6 | |
| --- | --- | --- | --- | --- | --- | --- | --- | --- | --- | --- |
| Treatment | Vehicle | TOP-N53<br>0.6 $\mu\text{g}$ | Vehicle | TOP-N53<br>1.21 $\mu\text{g}$ | Vehicle | TOP-N53<br>2.42 $\mu\text{g}$ | Vehicle | TOP-N53<br>4.84 $\mu\text{g}$ | Vehicle | TOP-N53<br>9.075 $\mu\text{g}$ |
| $\Delta\text{CVC}$ AUEC, Mean | 88.29 | 98.83 | 97.75 | 171.8 | 78.38 | 39.41 | 190.63 | 248.63 | 200.98 | 298.35 |
| $\Delta\text{CVC}$ AUEC, SD | 85.28 | 70.71 | 131.37 | 174.18 | 128.26 | 91.7 | 123.1 | 147.54 | 104.48 | 102.32 |

Arithmetic means with SD of individual ΔCVC AUEC (0-1440 min) computed from the time courses of ΔCVC are summarized per dose group and treatment in Table 3.

Arithmetic mean AUEC (0-1440 min) of CVC changes from baseline (ΔCVC AUEC) increased with increasing volumes except for Dose Group 3 for both vehicle and TOP-N53.

In the two highest dose groups, arithmetic mean ΔCVC AUEC were higher after administration of TOP-N53 versus vehicle, by 30.4 % in dose group 4 (800 µl) and by 48.4 % in dose group 5 (1500 µl) (Table 3). These differences were not statistically significant (non-parametric Wilcoxon’s matched pairs signed rank test). However, ΔCVC was an exploratory endpoint and the trial was not powered to detect significant differences for this outcome.

The time course of mean systemic arterial pressure (MAP) measured at the same time points as skin perfusion was not affected by increasing doses of TOP-N53 (Fig 4A, D, G, J, M) in line with the separate measurements of systolic and diastolic arterial pressure (Fig 3).

With the two highest subcutaneous dose volumes of 800 µl and 1500 µl injected (corresponding to 4.84 and 9.075 µg TOP-N53) a marked early (at ≤ 10 min) and delayed (at about 6 hours) increase in ΔCVC was observed with vehicle yet further augmented with TOP-N53. This response appeared related to the higher dosing volume as it was not or markedly less observed with the other dose groups (100, 200 and 400 µl volume) (Fig 4A, D, G, J, M). In fact, ΔCVC AUEC for vehicle was numerically higher in the 800 and 1500 µl dose groups compared to the 100, 200 and 400 µl dose groups but the differences were not significant (One way ANOVA with Tukey’s multiple comparison test).

Collectively, in this first-in-human trial in healthy male participants it was observed that single subcutaneous doses of TOP-N53 (4.84 µg or 9.075 µg) increased local skin perfusion around the injection site when compared to vehicle without affecting systemic arterial blood pressure in line with the design of the NO-releasing PDE5 inhibitor as a locally applied, locally acting drug and its expected locally confined vasoactive pharmacology. However, these effects were not statistically significant because of the low sample size with n=6 participants per group.

## Discussion

This first-in-human, randomized, double blind, vehicle controlled, single ascending dose, Phase I clinical trial establishes foundational evidence for the safety, tolerability, and pharmacodynamic activity of TOP-N53, a novel bifunctional molecule that integrates nitric oxide (NO) release with phosphodiesterase-5 (PDE5) inhibition. Administered as single ascending subcutaneous doses from 0.6 µg to 9.075 µg to healthy male volunteers, TOP-N53 demonstrated an excellent local and systemic safety and tolerability profile without relevant effects on systemic arterial blood pressure while at the same time, causing a locally confined increase in skin perfusion around the sites of subcutaneous administration.

The trial’s primary objective local safety and tolerability was met, as no untoward local reactions were observed (pain, erythema or redness, induration or oedema, haemorrhage or bruising, itching) at the injection site in any of the participants treated up to 24 h after SC injection of TOP-N53 or placebo (vehicle). Systemic safety and tolerability were equally favourable, with only minor, transient adverse events (e.g., dizziness and headache) and no relevant changes in closely monitored systemic arterial systolic, diastolic or mean blood pressure profiles up to a single subcutaneous dose of 9.075 µg TOP-N53 covering time periods from prior to until 7 days after the subcutaneous administration of study medication. Serious or severe adverse events did not occur in the trial (29).

Another key finding supporting the favourable systemic safety profile of SC administered TOP-N53 in this trial was the absence of meaningful systemic (plasma) exposure to TOP-N53 and its primary metabolite, TOP-52 at doses of TOP-N53 SC associated with local pharmacological efficacy reflected by the LSCI-verified increase in skin perfusion around the injection site. Indeed, in all participants, plausible values of plasma levels remained below the lower limit of quantification (LLOQ) at all doses of TOP-N53, corresponding to estimated free (unbound to plasma protein) plasma concentrations of < 0.06 pM for TOP-N53 and < 0.45 pM for TOP-52 considering the high plasma protein binding for TOP-N53 (99.97 %) and TOP-52 (99.79 %) These estimated maximum unbound plasma concentrations of TOP-N53 and TOP-52 were less than the minimum anticipated biological effect level (MABEL). MABEL was determined as 8 pM for TOP-N53 and 4 pM for TOP-52 corresponding to concentrations achieving 40 % relaxation in an isolated organ model of precontracted rat aortic rings with functionally intact endothelium (30) reflecting the highest potency for TOP-N53 and TOP-52 ever observed in non-clinical pharmacology studies.

TOP-N53 SC induced dose-dependent and sustained increases in local skin perfusion, as quantified by Laser Speckle Contrast Imaging (LSCI) as an exploratory endpoint of pharmacodynamic efficacy. Notably, trends towards increases in cutaneous vascular conductance (ΔCVC) were observed at doses of 4.84 and 9.075 µg TOP-N53 SC over vehicle SC. This increased skin perfusion indicates that locally, pharmacologically relevant target engagement was achieved that would result in an increase in cyclic guanosine monophosphate (cGMP) levels and subsequent activation of protein kinase G (PKG). Activation of PKG is well-known to induce vascular smooth muscle cell (VSMC) relaxation through several downstream mechanisms, including phosphorylation of IP3 receptor-associated cGMP kinase substrate (IRAG) (31, 32), resulting in inhibition of IP3-induced intracellular calcium release and activation of myosin light-chain (MLC) phosphatase (33), and inhibition of the RhoA/Rho-associated protein kinase (ROCK) pathway (34, 35) collectively contributing to sustained vasodilatory effects (21)

Subcutaneous injections of vehicle at the highest volumes of 800 µl in dose group 4 and 1500 µl in dose group 5 resulted in an initial (10 min after injection) and delayed (about 6 hours after injection) peak in ΔCVC from LSCI. The initial peak may be explained by the axon reflex (36): SC injections elicit antidromic activation of (nociceptive) sensory C-fibres accounting for release of vasoactive neuropeptides exemplified by substance P or calcitonin gene-related peptide (CGRP) from close axon terminals (that may be the same nerve endings harbouring both the transducer as well as the effector) causing rapid vasodilation hence, increased local skin perfusion. Increasing SC volumes may be expected to activate more C-fibres resulting in larger increases in skin blood flow. What caused the delayed ΔCVC remains an open question. This increase in skin perfusion could have been related to the higher injection volume, the excipients (Cremophore EL, PEG400) or both. For example, Cremophor EL has been described to activate mast cell degranulation (37) and subcutaneous injections may cause a delayed mast cell accumulation at the injection site (38, 39). Activated mast cells release histamine and synthetize prostanoids as PGD2, both may contribute to enhanced skin perfusion.

In the currently ongoing first-in-patients, first-on-wound clinical phase 2a trial with TOP-N53 (NCT06954597) a different, hydrogel-based formulation is used without Cremophor EL.

LSCI was selected for non-invasive measurements of skin blood flow in this trial due to its many advantages versus other methods as Laser Doppler Imaging (LDI) or Laser Doppler Flowmetry (LDF) such as superior spatial and temporal resolution, higher reproducibility, faster imaging (28, 40–43) and has previously been used to characterize skin perfusion following local and systemic vasodilators such as prostacyclin analogues or sildenafil (13, 44–46) including in patients with systemic sclerosis. Using LSCI, skin perfusion can be measured to a depth of approximately 300 μm. As a corollary LSCI mostly assesses perfusion of the superficial papillary plexus that include the nutritive papillary capillary loops (36).

While this first in human clinical trial with TOP-N53 successfully met its primary and secondary objectives, certain limitations merit discussion. The trial enrolled only healthy male volunteers of less than 56 years of age, in accordance with early-phase regulatory norms. Furthermore, the topical *on wound* route of administration envisaged for TOP-N53 in patients with cutaneous ulcerations was not feasible in healthy volunteers with intact skin. Representing a “worst case scenario” in terms of systemic exposure as it could happen from the later envisaged on wound administration, the sub-cutaneous route of administration was selected to enable a better and more translational assessment of systemic safety and tolerability risks than topical, *on skin* administration. However, this limits generalizability, particularly since chronic wounds affect a broad patient population across genders and comorbidities, often occur in the elderly and *on wound* administration is different from a subcutaneous injection. Future clinical trials include female participants and patient populations with active ulcers without age limits in compliance with current regulatory standards for inclusive trial design and on wound administration.

The trial does not address the question, whether the NO-releasing PDE5 inhibitor TOP-N53 outperforms a PDE5 inhibitor or other NO-donors (such as organic nitrates) on their own on skin perfusion. The non-smoking healthy volunteers enrolled in the trial are not expected being afflicted with endothelial dysfunction characterized by deficient endothelial NO such as in systemic sclerosis or diabetes mellitus who could experience additional benefit from TOP-N53 replenishing such depleted NO. On the other hand, in an earlier trial with healthy volunteers the PDE5 inhibitor sildenafil, administered orally potentiated local skin perfusion observed following topical administration of sodium nitroprusside, a NO donor, demonstrating pharmacodynamic amplification between PDE5 inhibitors and NO donors on skin blood flow (47).

The SC dosing by volume in this trial (from 100 µl in dose group 1 to 1500 µl in dose group 5 with identical volumes for vehicle and TOP-N53 within a dose group) precludes direct comparisons between dose groups, given that increasing volumes of vehicle resulted in increases in ΔCVC AUEC (0-1440 min) (illustrated by arithmetic means of ΔCVC AUEC in dose groups 4 and 5 compared to dose group 1). As indicated before, vehicle effects on skin perfusion may be caused by release of endogenous mediators. Whether the observed increase in skin perfusion by TOP-N53 in dose group 4 and 5 is explained by TOP-N53 alone or some pharmacological interaction of TOP-N53 with such endogenous mediators remains an open question.

Despite these limitations, this first in human trial with TOP-N53 lays a compelling foundation for Phase II clinical development in patients afflicted with chronic cutaneous wounds.

The systemic co-administration of oral PDE5 inhibitors and NO-donors (such as glycerol trinitrate; isosorbide mononitrate or dinitrate) is contraindicated because there is a risk of severe systemic blood pressure drops. For the first time, the current first in human trial delivered strong evidence, that subcutaneous administration of the locally applied, locally acting NO-releasing PDE5 inhibitor TOP-N53 fully dissociated local from systemic vasodilation in humans: While systemic arterial blood pressure was unaffected TOP-N53 increased skin perfusion locally confined around its application site. These findings herald that rigorously pursuing “*on wound* by design principles” as done in the discovery of TOP-N53 (25) enables to exploit the therapeutic benefits emanating from simultaneous NO-release to enhance cGMP synthesis and inhibition of its degradation by PDE5 for local treatments such as chronic cutaneous wounds, including digital ulcers in systemic sclerosis, diabetic foot ulcers and other conditions characterized by localized perfusion deficits.

Clinical findings with TOP-N53 in this first in human trial are strongly aligned with preclinical data, in which TOP-N53 improved cutaneous wound microcirculation attributed to pro-perfusion and also pro-angiogenic effects (26, 27). Given that chronic wounds, including diabetic, ischemic, and autoimmune-mediated ulcers often involve profound microvascular dysfunction, a potential ability of TOP-N53 to locally restore tissue perfusion without systemic side effects represents a potentially transformative therapeutic approach.

In conclusion, in this first in human clinical trial, the NO-releasing PDE5 inhibitor TOP-N53, administered as single, subcutaneous doses up to 9.075 µg integrated excellent local and systemic safety and tolerability with exhibiting the desired locally confined pharmacological activity to improve skin blood flow at the intended site of action. Of paramount importance at local pharmacological efficacy TOP-N53 did not affect systemic arterial blood pressure representing a strong differentiator from systemic vasodilators. A favourable pharmacokinetic profile encompassing high plasma protein binding that minimizes systemic exposure hence, risks largely accounts for the beneficial profile of TOP-N53 elaborated in the first in human trial. Based on these findings one may raise the notion that opposite to systemic vasodilators with high systemic plasma exposure yet presumably suboptimal exposure at chronic wounds, TOP-N53 as a locally, on wound applied, locally acting vasoactive compound would integrate optimal exposure at chronic wounds with minimal systemic exposure. If confirmed, this would translate into an improved benefit to risk ratio for TOP-N53 compared to systemic vasodilators. It remains to be seen, whether these findings translate to patients with chronic wounds, where TOP-N53 in a hydrogel formulation is administered topically. In this respect, a phase 2a trial with focus on safety and tolerability from a single topical *on wound* administration of TOP-N53 versus vehicle is currently recruiting patients with digital ulcers in systemic sclerosis (EUCTR 2024-511861-12-00; NCT06954597).

Later clinical trials will aim to determine the optimal therapeutic dose, explore repeated-dose safety, assess clinical endpoints such as wound closure and pain relief, and evaluate patient-reported outcomes. Additionally, biomarker-guided sub-trials may further elucidate the mechanistic underpinnings of TOP-N53’s dual activity and support precision-based application in vascular-compromised wound populations. The results of this first in human Phase I trial support the continued clinical development of TOP-N53 as a safe, locally acting vasoactive therapy with strong mechanistic and clinical potential for chronic wound indications.

## Data Availability

All data produced in the present study are available upon reasonable request to the authors

## Author contributions

Dr. Seitz, Dr. Ludin, Dr. Tenor, and Dr. Naef contributed to protocol development, data analysis, and manuscript preparation. Dr. Seitz was involved in the execution of the clinical trial. Prof. Cracowski contributed to the development of the LSCI methodology and critically reviewed the manuscript. Prof. Gerth critically reviewed the manuscript. Dr. Schaefer and Dr. Bhide contributed to the conceptualization of the manuscript and Dr. Bhide also coordinated manuscript preparation

## Acknowledgement

The contribution of the clinical operations team at CRS Clinical Research Services, Mannheim, Germany, is gratefully acknowledged. We also thank the study volunteers whose participation made this study possible.

